# The Association of the Neonatal Mortality Score and Clinical Outcomes in Newborns Delivered in a Tertiary Hospital in Cebu City, Philippines from December 2021 to July 2022

**DOI:** 10.1101/2025.06.24.25330161

**Authors:** Love Ellise G. Tarce, Lemuel Hope S. Galindo

**Affiliations:** Perpetual Succour Hospital of Cebu, Inc.; Cebu Institute of Medicine

**Keywords:** Neonatal mortality, mortality scoring system

## Abstract

“The Association of the Neonatal Mortality Score and Clinical Outcomes in Newborns Delivered in a Tertiary Hospital in Cebu City, Philippines from December 2021 to July 2022”

**Background:** Several neonatal scoring systems have been developed to predict illness severity and mortality risk. However, they are designed for high-income countries- using parameters that are not often measurable in some settings. The Neonatal Mortality Score was developed for resource-limited settings with the following parameters: level of consciousness, respiratory distress, gestational age, and birthweight.

**Objective:** To determine the association between the Neonatal Mortality Score and clinical outcomes among newborns

**Study design, setting and population:** Prospective cross-sectional study that included newborns delivered in a tertiary hospital in Cebu City, Philippines.

**Methods:** The mortality risk of neonates was assessed using the Neonatal Mortality Score. All were followed-up until discharge, and outcomes were recorded and analyzed.

**Results:** There was an association between the Neonatal Mortality Score and clinical outcomes with an AUC of 0.989 [95% CI, 0.971-1.000]. Using a cut-off of 13.5, the overall sensitivity was 100%, specificity was 97.43%, positive predictive value was 18.18%, and negative predictive value was 100%. Among the parameters used, respiratory distress, and low birthweight showed an association with clinical outcomes. The admission disposition and length of stay also showed a significant association with Neonatal Mortality Scores.

**Conclusion:** The Neonatal Mortality Score may be used in resource-limited settings to anticipate neonatal mortality, and predict admission disposition and length of stay, stratifying neonates with increased risk for death, thereby reducing early complications.

## BACKGROUND OF THE STUDY

The neonatal period, defined as the first 28 days of life, is the most vulnerable time for a child’s survival. With most deaths under the age of five occurring during this period, (1) neonatal mortality is a reflection of a country’s socio-economic status, maternal health service utilization and overall health care quality.

In the Philippines, a total of 11, 948 neonatal deaths were recorded in 2018, with 7.2 neonatal deaths per 1000 live births. Out of these, 44.6% occurred within the first seven days of life with bacterial sepsis and respiratory distress of the newborn as the leading causes of mortality in this age group. (1) With a majority of all newborn deaths from preventable and treatable causes such as complications related to prematurity, sepsis, and birth asphyxia (2), improving the quality of care will save the most lives, but this requires the availability of essential commodities and proper education and training of healthcare workers.

Another challenge facing healthcare facilities is the prolonged hospital length of stay (LOS) of neonates. This has important implications for health outcomes, quality of care and patient safety. Decreasing the patient’s length of stay can more effectively allocate resource consumption of the facilities and decrease the economic and social burden of the patient’s family. (3) Predictors of increased hospital length of stay in neonates admitted in the NICU were low birth weight, those less than 37 weeks gestational age and developing hospital acquired infection 48 hours after admission. (4)

The transition from fetus to neonate entails a series of rapid and dramatic physiologic changes. Nevertheless, approximately 90% of births transition successfully without any or only limited assistance. (5) This entails successful transition in the (a) respiratory, (b) cardiovascular, and (c) metabolic aspects. (6) It was concluded that neonates may take up to 30 minutes to establish a stable functional residual capacity and may need up to 90 minutes to establish a stable and regular respiratory rate and pattern. (7) (8) Immediate cardiovascular transition appears to be completed around 1 hour of life, with functional closure of fetal circulatory shunts continuing for 24-48 hours of life. (9) Lastly, a successful metabolic transition to extrauterine life may be indicated if neonates are without clinical manifestations of hypoglycemia by 1-3 hours after birth, although further observation is needed. (10) Thus, it was concluded that the successful immediate transition of a neonate to extrauterine life is completed within 1-3 hours of life. (6)

Failure of the neonate to successfully transition may be due to compounding factors, which may contribute to neonatal mortality. Among these are prematurity, sepsis, low Apgar scores, low birthweight, birth asphyxia, low socioeconomic status, congenital malformations, and cesarean section delivery (11) (12) (13) (14) (15) (16). From these factors, several neonatal scoring systems have been developed to identify high-risk neonates with poor prognoses or those who are suitable for a particular intervention. These early warning scores assign a certain value to maternal and neonatal parameters to derive a composite score to identify high-risk neonates.

The Score for Neonatal Acute Physiology (SNAP) (17) model developed in 1989-1990 evaluates 28 variables that are to be assessed within the first 24 hours of admission. This was further improved using the Score for Neonatal Acute Physiology- Perinatal Extension (SNAP-PE), with birthweight, small for gestational age, and low Apgar score on the 5th minute of life as additional parameters. Updated SNAP II and SNAP-PE II were then published in 2001, and use measurable parameters that vary with illness severity, such as serum pH and PO2/FiO2 ratio. (18) In 1993, the Clinical Risk Index for Babies (CRIB) score (19) was established and utilizes six parameters to be considered during the first 12 hours of life but is only used for neonates less than 32 weeks of gestation and/or birthweight less than or equal to 1500 grams. This was then updated in 2003 by Parry et al. (20). Furthermore, the Neonatal Therapeutic Intervention Scoring System (NTISS) uses the intensity of treatment and resources used, such as respiratory therapies, drug therapies and monitoring, to calculate and predict illness severity. (21)

Notably, these scoring systems were primarily developed for high-income countries. The local health centers and birthing centers cannot utilize the aforementioned tools, as they include laboratory-derived and treatment-derived parameters that are not often available or measurable in resource-limited settings.

According to Fleisher et al., (22) important features of a neonatal scoring system include (a) ease of use; (b) its applicability early in the course of hospitalization; (c) being able to predict mortality, specific morbidities, or cost for various categories of neonates; and (d) its usefulness for all groups of neonates to be described.

To improve the quality of neonatal care in the Philippines, especially in resource-limited settings, a predictive scoring system that is feasible for routine use is needed to aid healthcare providers in objectively assessing the newborn mortality risk for early referral to a specialist and/or immediate transfer to a more equipped facility.

### Neonatal Mortality Score

In 2019, the Neonatal Mortality Score (NMS) was derived and validated as a prognostic score for neonatal mortality for neonates admitted to the Neonatal Intensive Care Unit (NICU) in Ethiopia. (23) The total Neonatal Mortality Score ranges from 0 to 52, with a predicted mortality of 4% and 100% respectively. A score of 12 corresponds to 50% probability of mortality. With a sensitivity of 81%, a specificity of 80%, a positive predictive value of 58%, and a negative predictive value of 83%, this can be a useful screening tool for neonatal mortality upon admission. The parameters included were the following: level of consciousness, respiratory distress, gestational age, and the birthweight of the neonate. The said parameters were measurable and feasible, thus, it was considered to determine the applicability of the Neonatal Mortality Score in our setting.

## OBJECTIVES

### General

This study aimed to determine the association between the Neonatal Mortality Score and clinical outcomes among newborns delivered in a Tertiary Hospital in Cebu City, Philippines from December 2021 to July 2022.

### Specific

1. To determine and compare the demographic and clinical characteristics of the neonates delivered from December 2021 to July 2022 as to the following:

a. Gestational age, weeks
b. Birthweight, grams
c. Sex
d. Mode of delivery
e. Comorbidities
f. Admission disposition (Level I, Level II, Level III)
g. Length of hospital stay (short, medium or long)
h. Outcome (alive or died)
2. To determine the frequencies of the scores for each parameter of Neonatal Mortality Score on admission among the neonates who were discharged alive or neonates who died as follows:

a. Level of consciousness
b. Respiratory distress
c. Gestational age, weeks
d. Birthweight, grams
3. To determine and compare the relationship between the Neonatal Mortality Score and the length of hospital stay among neonates who were discharged alive or those who died.
4. To determine and compare the relationship between the Neonatal Mortality Score and the admission disposition (Level I, Special Care Unit, Neonatal Intensive Care Unit) among those who were discharged alive or those who died.
5. To determine and compare the relationship between the Neonatal Mortality Score and the clinical outcome among those alive and those who died.

## RESEARCH METHODOLOGY

### STUDY DESIGN

This was a facility-based prospective cross-sectional study.

### STUDY SETTING

This study was conducted at a tertiary hospital in Cebu City, Philippines.

### STUDY POPULATION

#### Inclusion criteria

Data of neonates delivered in a tertiary hospital from December 2021 to July 2022 were used.

#### Exclusion criteria

a) Neonates who died before the 3^rd^ hour of life
b) Neonates who were discharged against medical advice
c) Stillbirth

### SAMPLE SIZE CALCULATION AND SAMPLING PROCEDURE

The sample population of this study was neonates delivered in a tertiary hospital in Cebu City, Philippines from December 2021 to July 2022. Sample size and power computation were done and are specified below.

| Sample Size for Frequency in a Population |  |
| --- | --- |
| Population size(for finite population correction factor or fpc)($N$): | 868 |
| Hypothesized % frequency of outcome factor in the population ($p$): | 60%±5 |
| Confidence limits as % of 100(absolute ± %)($d$): | 5% |
| Design effect (for cluster surveys-$DEFF$): | 1 |
| Sample Size ($n$) for 95% Confidence Level | |
| Confidence Level | Sample Size |
| 95% | 260 |

| Equation |  |
| --- | --- |
| Sample size $n = [DEFF * Np(1-p)] / [(d^2 / Z_{1-\alpha/2}^2 * (N-1) + p * (1-p)]$ | |
| Results from OpenEpi, Version 3, open source calculator--SSPropor |  |

Using the OpenEpi version 3 sample size calculator, with the estimated true proportion of 60% of neonatal admissions from 2019-2020 in the institution, the sample size required was 260 subjects with a desired precision of estimate of 0.05 and 0.95 confidence interval. Total enumeration was used based on the characteristic population of interest and objective of the study, with a total of 352 subjects.

### DATA COLLECTION PROCEDURE

Prior to data collection, all pediatric residents who rotated in the nursery were oriented as to the study’s objectives and the data collection tool that was used. The Neonatal Mortality Score was discussed as well as the evaluation of the neonates and classifying them under certain parameters.

Neonates delivered from December 2021 to July 2022 were included in the study. Initial data such as the demographic and clinical profile (gestational age, birthweight, sex, mode of delivery) were collected. On the third hour of life, the neonate’s mental status and respiratory distress classification were assessed. The total Neonatal Mortality Score from the four parameters was documented. The neonates were then followed-up until discharge and other data such as the comorbidities and other diagnoses, admission disposition, length of hospital stay, and outcomes (alive/died) were recorded in the data collection tool. The said data were then logged in Microsoft Excel using coded numbers. For confidentiality, no copies of any portion of the patients’ records such as photos or screenshots were taken.

#### Neonatal Mortality Score

The Neonatal Mortality Score was developed and validated by Mediratta et al. in 2019 using four variables to predict neonatal outcome upon admission using the following parameters: level of consciousness, respiratory distress, birthweight, and gestational age. (23) Under level consciousness, neonates were classified as alert, irritable, lethargic and comatose, with scores of 0, 6, 11, and 16, respectively. For respiratory distress, neonates were assessed as having none, mild, moderate and severe distress, with scores of 0, 3, 11, and 14, respectively. Patients were also scored according to gestational age, with greater than or equal to 37 weeks with a score of 0, those delivered at 32-36 weeks with a score of 1, and those born less than 32 weeks with a score of 10. For the neonate’s birthweight, those weighing more than or equal to 2500 grams were given a score of 0, 1500-2400 grams were given a score of 5, and those less than 1500 grams were given a score of 12. The scores for each parameter were added and the total Neonatal Mortality Score ranged from 0 to 52.

### DATA ANALYSIS

Data was entered in Microsoft Excel Spreadsheet and coded where necessary. SPPS version 22.0 was used for data analysis.

Frequency distribution and percentage were used to present several categorical data under the demographic and clinical characteristics of patients as well as their initial Neonatal Mortality Score, and outcomes.

Descriptive statistics such as Mean, Standard Deviation (SD), and Minimum and Maximum values were reported to describe the numerical variables under the demographic and clinical characteristics of the patients. The same was also used to present the initial Neonatal Mortality Score of the patients.

Testing for normality was done, and a comparison between neonates discharged alive and died was performed using the Mann-Whitney test since the data were not normally distributed. For the comparison between NMS and admission disposition and between the length of stay, the Kruskal Wallis test was used. The power of the Neonatal Mortality Score to predict neonatal mortality was evaluated by means of the Receiver Operating Characteristics (ROC) curve. Sensitivity, specificity, positive and negative predictive values were determined and calculated for different cut-off scores.

A p-value of less than 0.05 was considered statistically significant for all tests.

## RESULTS

### Clinico-demographic profile of the neonates

A total of 380 neonates were delivered in the nursery during the study period (December 2021 to July 2022). Twenty-eight neonates were then excluded from the study. Among the 352 neonates who were enrolled in the study, the majority were delivered via normal spontaneous delivery (66.8%). The number of males and females was almost equal, comprising 50.6% and 48.9% of the population, respectively. Only 2.3% had comorbidities such as congenital heart disease and congenital anomalies. Delivered neonates were 27 weeks to 41 weeks of gestation, with an average birthweight of 2870 grams (SD= 477.08). For admission disposition, 4.8% were admitted in the NICU, 24.1% were admitted in level II and the rest (71.0%) were well-babies and admitted to level I. The mean length of hospital stay was 5 days (SD= 6.08). Out of the neonatal deliveries, 350 (99.4%) were discharged alive and only 2 (0.6%) died during the hospital stay. The general characteristics of the neonates delivered in the nursery are shown in Table 1.

**Table 1.**
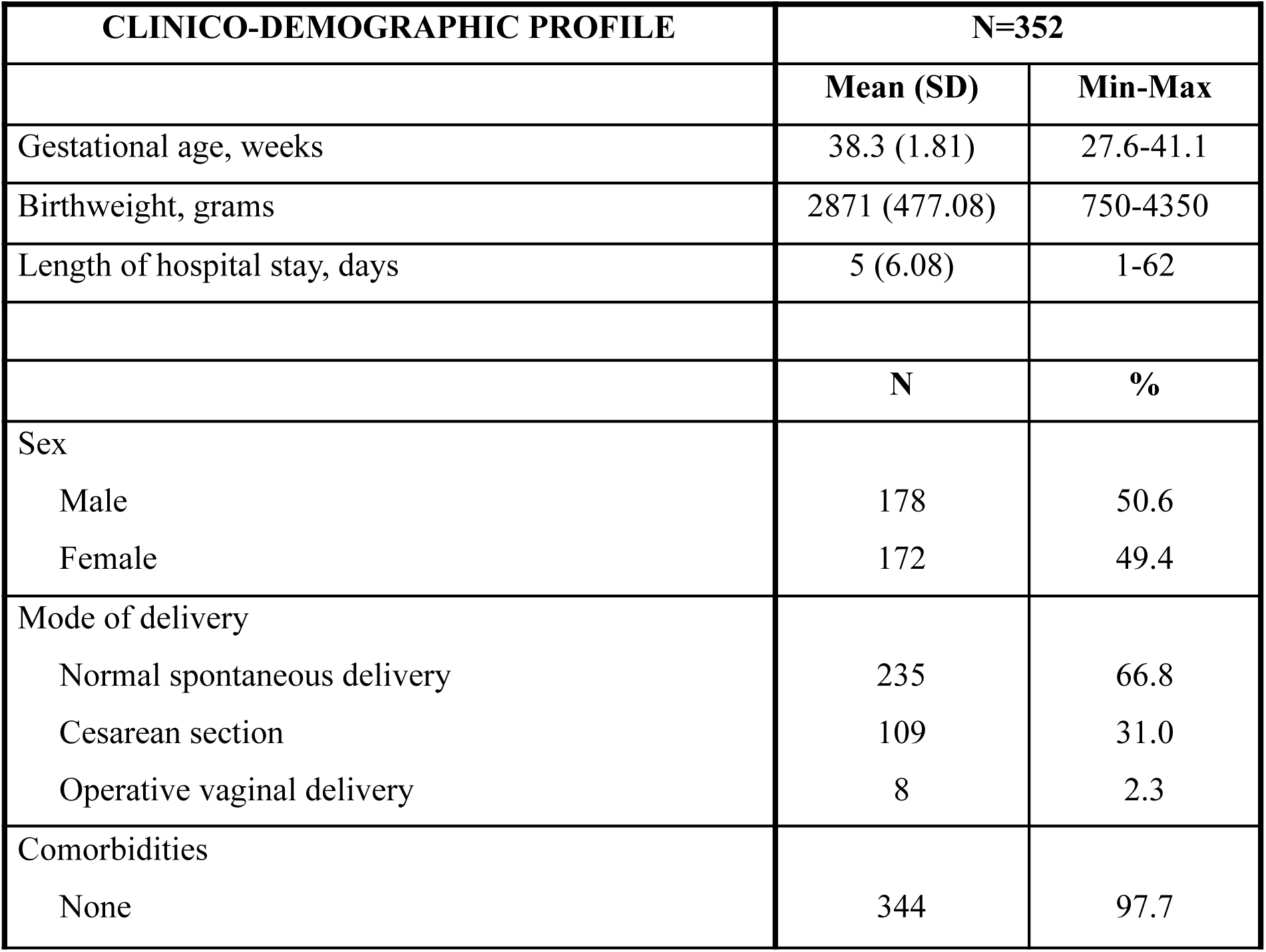
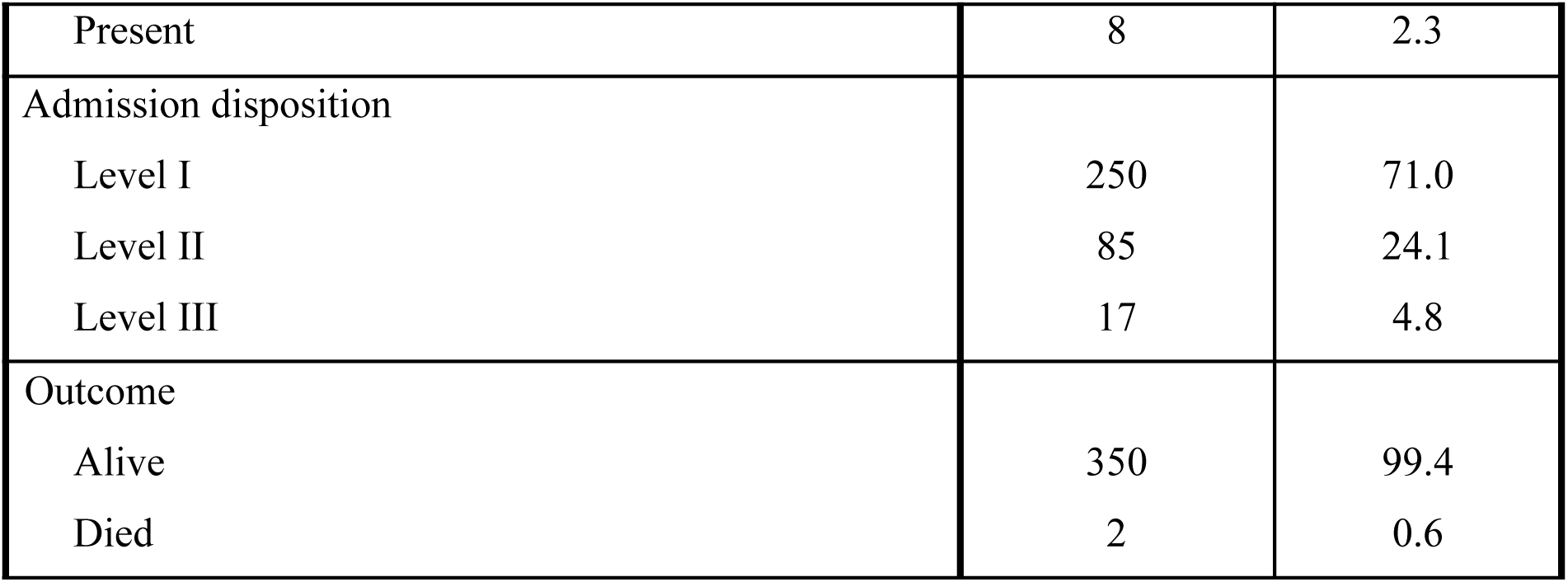
General Characteristics of Neonatal Deliveries.

| CLINICO-DEMOGRAPHIC PROFILE |  | N=352 |  |
| --- | --- | --- | --- |
|  |  | Mean (SD) | Min-Max |
| Gestational age, weeks |  | 38.3 (1.81) | 27.6-41.1 |
| Birthweight, grams |  | 2871 (477.08) | 750-4350 |
| Length of hospital stay, days |  | 5 (6.08) | 1-62 |
|  |  | N | % |
| Sex |  |  |  |
| Male |  | 178 | 50.6 |
| Female |  | 172 | 49.4 |
| Mode of delivery |  |  |  |
| Normal spontaneous delivery |  | 235 | 66.8 |
| Cesarean section |  | 109 | 31.0 |
| Operative vaginal delivery |  | 8 | 2.3 |
| Comorbidities |  |  |  |
| None |  | 344 | 97.7 |

|  |  |  |
| --- | --- | --- |
| Present | 8 | 2.3 |
| Admission disposition |  |  |
| Level I | 250 | 71.0 |
| Level II | 85 | 24.1 |
| Level III | 17 | 4.8 |
| Outcome |  |  |
| Alive | 350 | 99.4 |
| Died | 2 | 0.6 |

The Neonatal Mortality Score comprises four parameters- the level of consciousness, respiratory distress, gestational age, and birthweight. As shown in Table 2, under the level of consciousness, all neonates were alert upon assessment. Sixteen percent presented with a degree of respiratory distress, which included the neonates who died. From the study population, 50% who died were more than 37 weeks of gestation, and the remaining 50% who died were less than 37 weeks of gestation. All neonates who died weighed less than 2500 grams.

**Table 2.** Neonatal Mortality Score Parameters among Neonates Discharged Alive and Died.

| PARAMETERS | ALIVE<br>(N=350) |  | DIED<br>(N=2) |  | p-value |
| --- | --- | --- | --- | --- | --- |
|  | n | % | n | % |  |
| Level of consciousness |  |  |  |  | NA |
| Alert | 350 | 99.4 | 2 | 100 |  |
| Respiratory distress |  |  |  |  | 0.025 |
| None | 296 | 84.6 | 0 | 0 |  |
| Present | 54 | 15.4 | 2 | 100 |  |
| Gestational age |  |  |  |  | 0.158 |
| ≥37 weeks | 322 | 92.0 | 1 | 50 |  |
| <37 weeks | 28 | 8.0 | 1 | 50 |  |
| Birthweight |  |  |  |  |  |
| ≥2500 grams | 294 | 84.0 | 0 | 0 | 0.027 |
| <2500 grams | 56 | 16.0 | 2 | 100 |  |

*p-value was calculated using Chi-square test*

With a p-value less than 0.05 for both the respiratory distress and birthweight parameter, there was a significant relationship between the presence of respiratory distress and birthweight of <2500 grams on the clinical outcome of neonates.

#### Admission disposition

There was a significant association between the admission disposition and the Neonatal Mortality Score with a p-value of 0.00 (Table 3).

**Table 3.** Admission Disposition and the Neonatal Mortality Score of Newborns Discharged Alive and Who Died.

| Admission Disposition | Neonatal Mortality Score |  |
| --- | --- | --- |
|  | Mean (SD) | p-value |
| Level I | 0.70 (1.868) | 0.000 |
| Level II- SCU | 3.38 (4.590) |  |
| Level III- NICU | 13.00 (8.916) |  |

*p-value was calculated using the Kruskal Wallis test*

#### Length of stay

There was also a significant association between the length of stay and the Neonatal Mortality Score with a p-value of 0.00 (Table 4).

**Table 4.** Length of Stay and The Neonatal Mortality Score of Newborns.

| Length of Stay | Neonatal Mortality Score |  |
| --- | --- | --- |
|  | Mean (SD) | p-value |
| Short stay (≤ 5 days) | 0.87 (2.101) | 0.000 |
| Medium stay (6-10 days) | 4.06 (4.850) |  |
| Long stays (>10 days) | 13.06 (8.740) |  |

*p-value was calculated using the Kruskal Wallis test*

#### Clinical outcomes

The clinical outcomes as to alive and died also showed a significant association with the Neonatal Mortality Score with a p-value of 0.002 (Table 5).

**Table 5.** Clinical Outcomes and the Neonatal Mortality Score.

| Clinical Outcome | Neonatal Mortality Score |  |
| --- | --- | --- |
|  | Mean (SD) | p-value |
| Alive | 1.81 (3.9) | 0.002 |
| Died | 24.5 (12.0) |  |

*p-value was calculated using the Mann Whitney test*

Plotting the values in the ROC curve, the Area under the curve (AUC) was 0.989 [95% CI, 0.971-1.000] as shown in Figure 2, which indicated that the Neonatal Mortality score was an excellent tool used as a predictor for mortality.

**Fig. 2.**
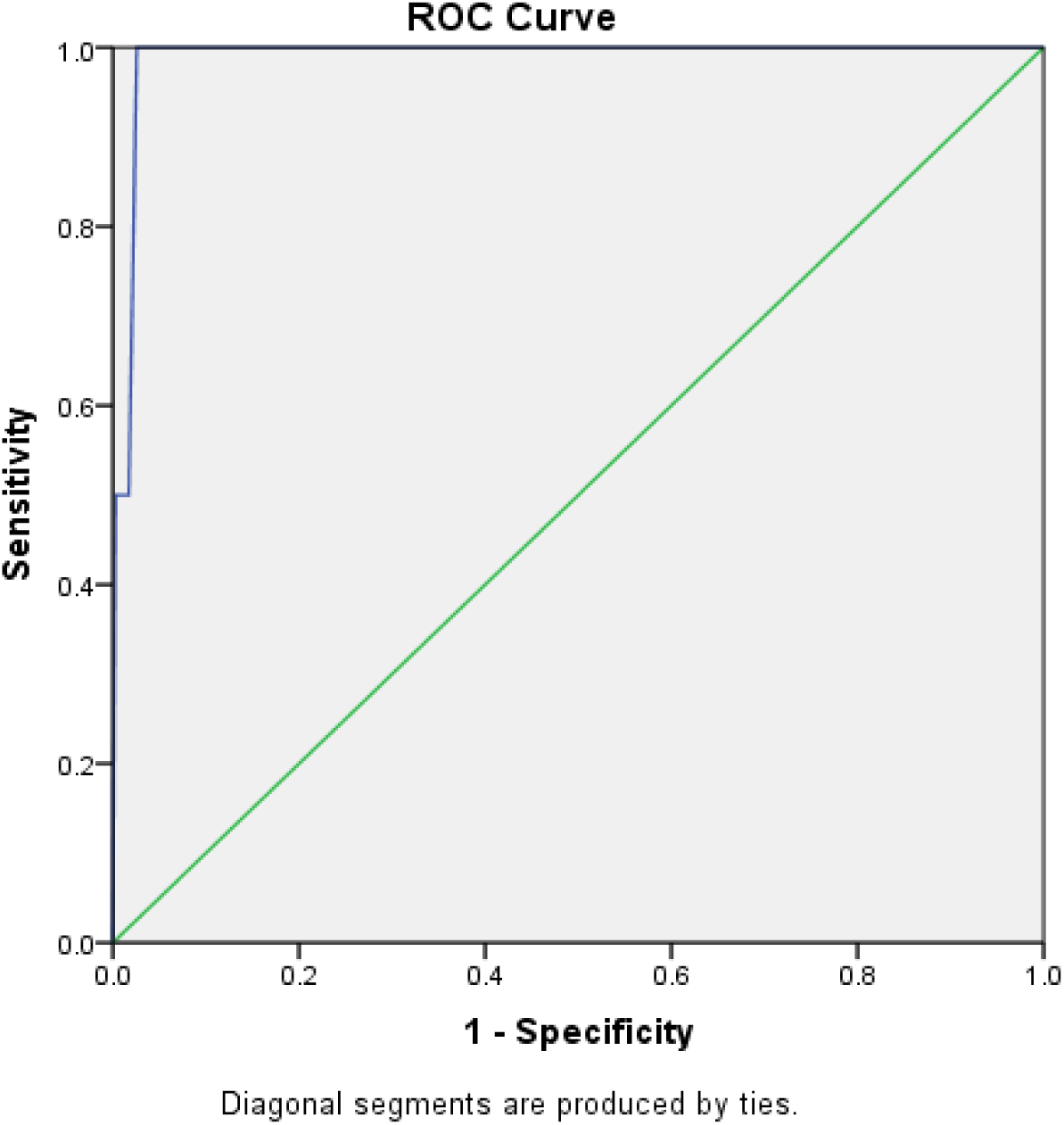
Receiver Operating Characteristics (ROC) Curve for Neonatal Mortality Score for Prediction of Mortality.

Using the cut-off Neonatal Mortality Score of 13.5 in estimating the overall mortality, results showed a sensitivity of 100%, specificity of 97.43%, positive predictive values of 18.18%, a negative predictive value of 100%, and an accuracy of 97.44% as shown in Table 6.

**Table 6.** Neonatal Mortality Score and Clinical Outcome Crosstabulation.

|  |  |  | OUTCOME |  | Total |
| --- | --- | --- | --- | --- | --- |
|  |  |  | Alive | Died |  |
| NMS<br>SCORE | Alive | Count | 341 | 0 | 341 |
|  |  | % within NMS SCORE | 100.0% | 0.0% | 100.0% |
|  |  | % within OUTCOME | 97.4% | 0.0% | 96.9% |
|  | Died | Count | 9 | 2 | 11 |
|  |  | % within NMS SCORE | 81.8% | 18.2% | 100.0% |
|  |  | % within OUTCOME | 2.6% | 100.0% | 3.1% |
| Total |  | Count | 350 | 2 | 352 |
|  |  | % within NMS SCORE | 99.4% | .6% | 100.0% |
|  |  | % within OUTCOME | 100.0% | 100.0% | 100.0% |

## DISCUSSION

This study evaluated the association between the Neonatal Mortality Score (NMS) and the clinical outcome of neonates delivered in a tertiary nursery in Cebu City, Philippines. Overall, only 0.6% of neonates died during the study duration. The present study documented that the NMS of neonates who died in the nursery was higher than those who were discharged alive. A Neonatal Mortality Score of ≥ 13.5 predicted overall mortality with a sensitivity of 100%, specificity of 97.43%, positive predictive value of 18.18%, and a negative predictive value of 100%.

This result supported the study done by the original author Mediratta et al. (23) in which AUC was 0.85 (95%CI 0.82-0.89), compared to the AUC in this study which was 0.989 (95% CI, 0.971-1.000). Similarly, the NMS showed excellent negative predictive value but had poor positive predictive value. In this study, a major factor for this was that the number of neonatal mortalities for the study duration was decreased. The predictive value depends on the sensitivity and specificity of a test and the prevalence, and when the prevalence of neonatal mortality is low, the positive predictive value may also be decreased, despite having high sensitivity and specificity. Higher NMS scores- neonates predicted to have increased risk for mortality- in those who were eventually discharged alive may be due to other confounding factors such as immediate resuscitation, availability of resources, and timely management during the admission.

Of the four parameters in the NMS, there was a significant association between a birthweight of less than 2500 grams and the clinical outcome, and between the presence of respiratory distress and the clinical outcome. Birthweight is a common physiologic parameter included in other mortality risk assessment tools in newborns such as CRIB (19), CRIB II (24), SNAP (17), SNAP II and SNAP-PE (18) and has long been implicated as a major risk factor for neonatal mortality. This is attributed to the newborn’s inability to successfully physiologically transition to the extrauterine environment compared to those with a normal birthweight. (25) Decreased birthweight has been associated with hypoglycemia, hypothermia, electrolyte imbalance, infection and difficulties in feeding, predisposing this group of neonates to mortality within the first 28 days of life. (26)

The presence of respiratory distress, on the other hand, is an uncommon parameter in scoring systems that was included in the NMS. This can be due to the fact that the other scoring systems measured the degree of respiratory distress through laboratory work-up such as arterial blood gases, or the need for a certain oxygen levels for the patient to be stable. The association between respiratory distress and clinical outcome supports the study done by Swarnkar et al. (27) that neonates with respiratory distress are two to four times more likely to die than those without respiratory distress.

Gestational age is a common parameter in the previously mentioned neonatal mortality scoring systems. Problems associated with prematurity are mainly due to the underdevelopment of organ systems and include respiratory distress, apnea, intraventricular hemorrhage, cardiac dysfunction, sepsis, anemia, metabolic dysfunction, and hypothermia. (28) With this, gestational age has been associated with an increased risk for mortality, with approximately two-thirds of preterm neonates dying within the first 72 hours of admission (29). As opposed to this study, decreasing gestational age did not have a significant association with clinical outcomes. Factors that may have contributed to this were comorbidities, perinatal asphyxia, infection or other risk factors of the patients- the presence of which was not included in the study assessment.

It was not possible to determine if there was an association between the level of consciousness and the clinical outcome since all neonates were alert at the time of assessment. On the other hand, in the original NMS study (23), altered mental status and respiratory distress had increased strength of association compared to that of low birthweight and prematurity. Sepsis, the leading cause of neonatal morbidity, may present with altered mental status along with other physiologic abnormalities. (30) In the absence of resources for confirming the diagnosis of sepsis through laboratory tests, clinical suspicion is key to early treatment.

There was also a significant association between the Neonatal Mortality Score and the admission disposition, where neonates who had higher NMS were admitted in Level III. This study also demonstrated a significant association between the NMS and the length of hospital stay. The aforementioned secondary outcomes were not measured in the original study, but may be of help in areas with limited-resource settings to facilitate immediate referral to an institution much more capable to handle more complex cases requiring certain interventions (i.e. umbilical vein catheter insertion, antibiotics, continuous positive airway pressure or mechanical ventilation). In addition, the use of limited medical equipment may be prioritized for patients with higher NMS scores.

## CONCLUSION

The Neonatal Mortality Score may be used to anticipate neonatal mortality in a limited-resource clinical setting, with good sensitivity, specificity and negative predictive value. This allows members of the healthcare team to quickly identify neonates in need of additional interventions. In this study, neonatal mortality was associated with decreased birthweight and the presence of respiratory distress.

The aforementioned scoring system can also be utilized to predict admission disposition, prioritizing the use of limited resources and equipment for at-risk neonates identified by the score. Moreover, increased length of hospital stay was also associated with an increased score, enabling the efficient use of personnel and resources.

Therefore, this mortality scoring system can help in risk-stratifying neonates and identifying those with increased risk for death, providing an opportunity to reduce early neonatal complications, and consequently, enhancing overall patient care.

## RECOMMENDATIONS

This study was done in a tertiary hospital in Cebu City, Philippines. It is recommended that further study using the Neonatal Mortality Score be done in another resource-limited setting before introducing it as a clinical or public health tool. Moreover, concomitant diagnoses or management can affect the results of this study and is therefore recommended to be included in the analysis of data of the neonates fulfilling the inclusion criteria. With the decreased number of mortalities, it is also strongly recommended to increase the duration of the study to ensure that the same findings may be concluded with more mortalities included in the study population. Further, the NMS was assessed only at the third hour of life- the time of successful transition of a neonate to extrauterine life. The authors recommend that a repeat assessment be done at 24 hours of life to account for symptoms that may develop at a later time which may have a significant effect on the clinical outcome. It is also therefore recommended to determine the applicability of this tool and compare the scores between the third hour of life and at 24 hours of life

## Data Availability

All data produced in the present work are contained in the manuscript.

## APPENDICES

## APPENDIX A NEONATAL MORTALITY SCORE

NEONATAL MORTALITY SCORE
| PARAMETERS | SCORE |
| --- | --- |
| Level of Consciousness<br>▪ Alert<br>▪ Irritable<br>▪ Lethargic<br>▪ Comatose | 0<br>6<br>11<br>16 |
| Respiratory Distress<br>▪ None<br>▪ Mild<br>▪ Moderate<br>▪ Severe | 0<br>3<br>11<br>14 |
| Gestational age, weeks<br>▪ $\geq 37$<br>▪ 32-36<br>▪ $< 32$ | 0<br>1<br>10 |
| Birthweight, grams<br>▪ $\geq 2500$ | 0<br>5 |
| ▪ 1500-2499<br>▪ <1500 | 12 |
| TOTAL |  |

## APPENDIX B DATA COLLECTION TOOL

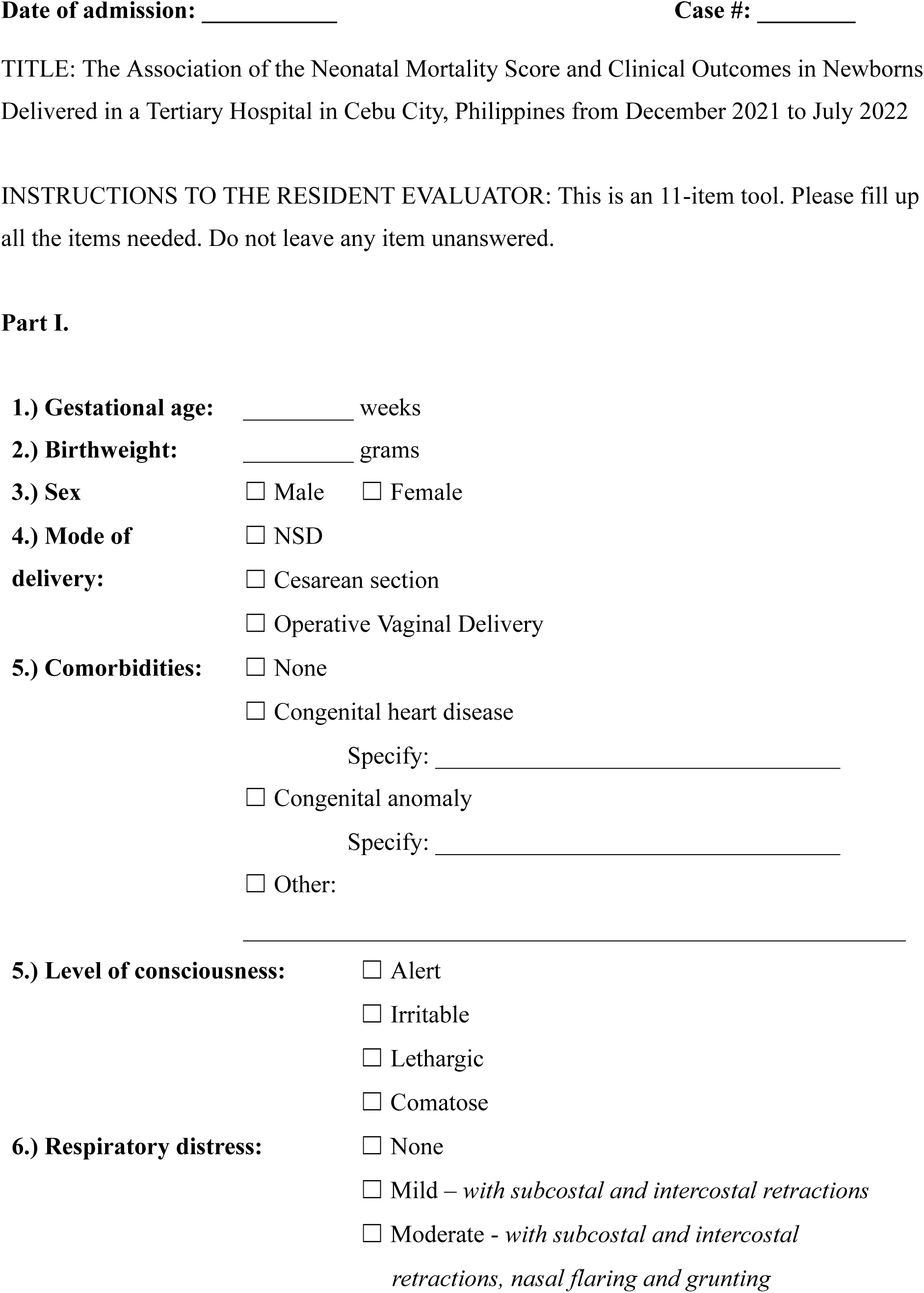
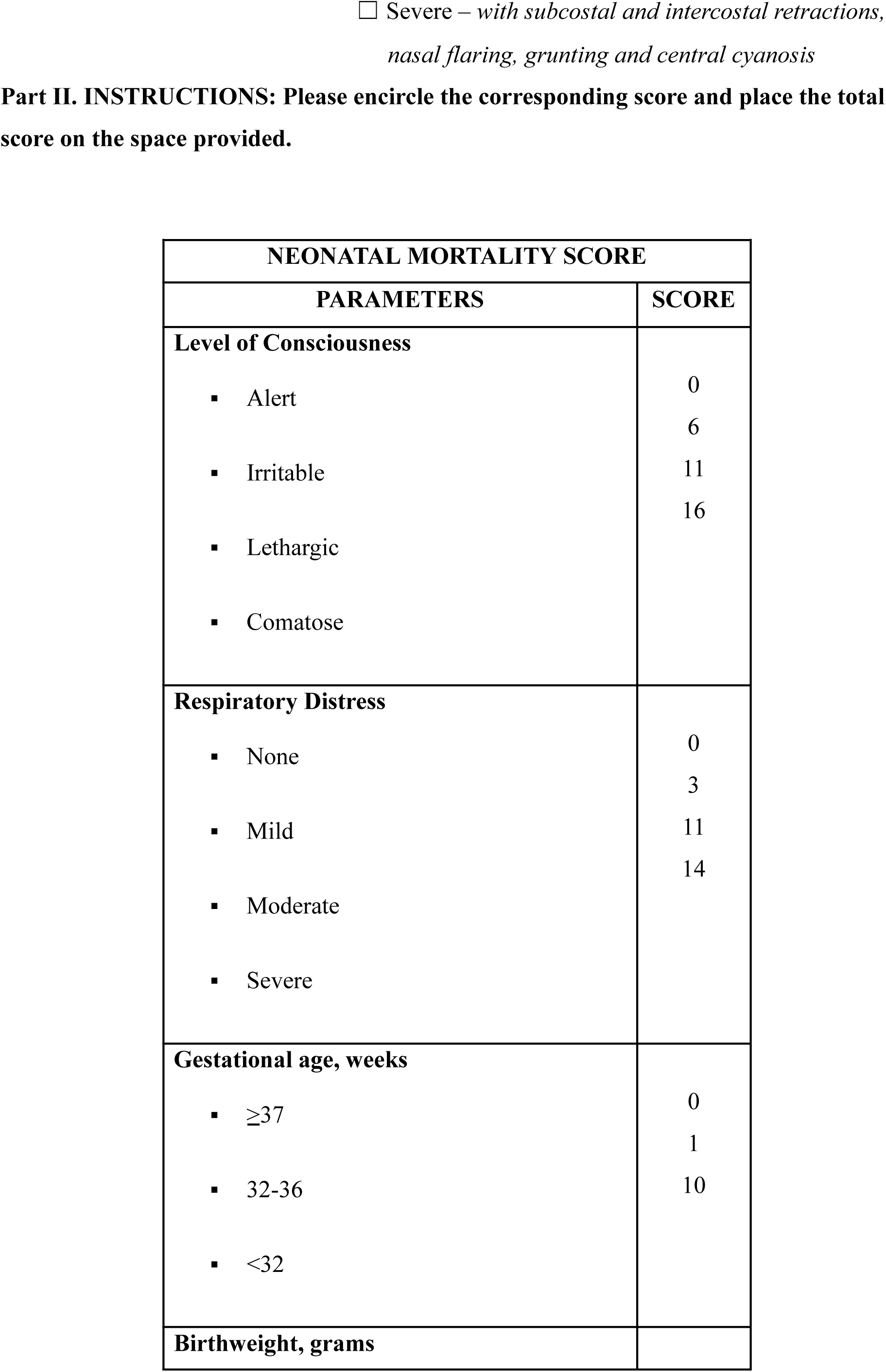
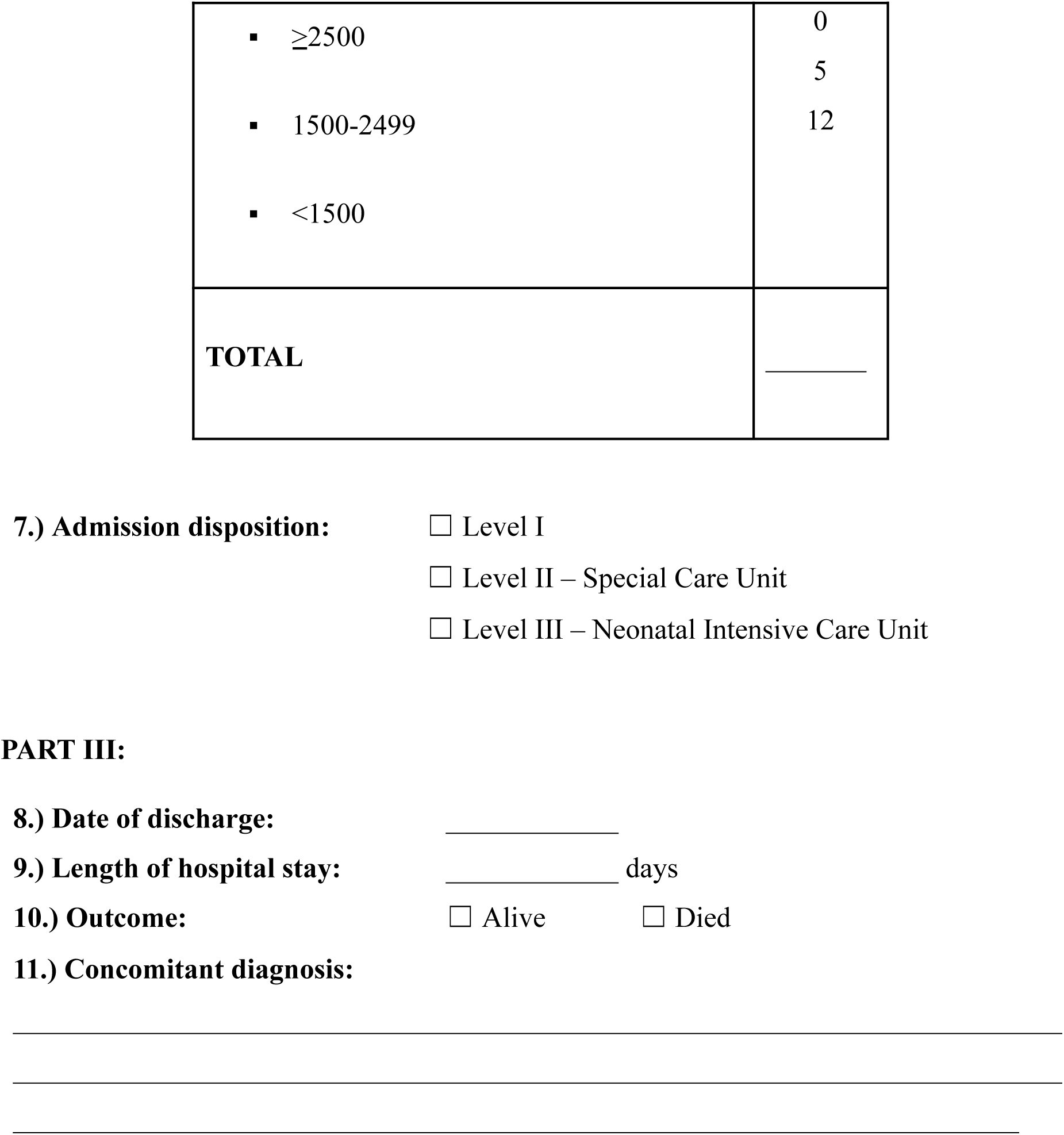

## APPENDIX C INFORMED CONSENT

### Informed Consent Form for the research

Date:________________

#### Introduction and Purpose

Before you consent for your newborn to be part of this study, it is important that you understand why this research is being done and how this will be conducted. Please read the following information carefully.

A number of neonatal disease scoring systems have been developed to predict neonatal illness severity and risk for mortality. The Neonatal Mortality Score was developed primarily for resource-limited settings and uses the following parameters upon admission: level of consciousness, respiratory distress, gestational age, and birthweight. The score has excellent discrimination and has been validated as a tool to predict mortality, useful for risk-stratifying and allowing proper resource allocation.

This study aims to determine the association of the Neonatal Mortality Score and clinical outcome among newborns delivered in a tertiary hospital from December 2021- July 2022.

Please ask the researcher if there is anything that is not clear or if you need more information on something.

#### Selection of Participants

This study will use data of newborns delivered in a tertiary hospital from December 2021 to July 2022

#### Procedure & Duration

The newborn will be assessed at the nursery on the third hour of life using the Neonatal Mortality Score with the following parameters: level of consciousness, respiratory distress, gestational age, and birthweight. The findings will be recorded, as well as the newborn’s outcome upon discharge.

#### Risks & Discomforts

The newborn will be assessed as part of the routine physical examination by a trained healthcare professional. Rest assured, non invasive procedures will be done in relation to the study. If you have any concerns, you may inform the researcher immediately.

#### Benefits

The researcher hopes that this study can help determine the association of the Neonatal Mortality Score and the clinical outcome among newborns. This can help in the said score’s applicability in low-resource settings, improving timely identification of neonates in need of additional interventions, and allow for proper allocation of personnel and resources.

#### Confidentiality

Strict confidentiality will be observed during the entire process of the data collection in which there will be no patient identifier. The data will then be logged in Microsoft Excel using coded numbers. The file will be protected with the author’s password and no other person will have knowledge of this password. We will not tell other people that you are part of this research and we will not share information about your newborn to anyone who does not work in the research study.

#### Voluntary Participation

Your consent for your newborn’s inclusion in this study is voluntary. If you choose to consent, you will be asked to sign a consent form. After you sign the consent form, you are still free to withdraw at any time and without giving a reason. Withdrawing from this study will not affect the relationship you have, if any, with the researcher. If you withdraw from the study before data collection is completed, the data collected will be deleted and will not be included in the study.

#### Certificate of Consent

I have read and understood the provided information. I have had the opportunity to ask questions about it and any questions that I have asked have been answered to my satisfaction. I consent voluntarily for my child to be a participant in this study.

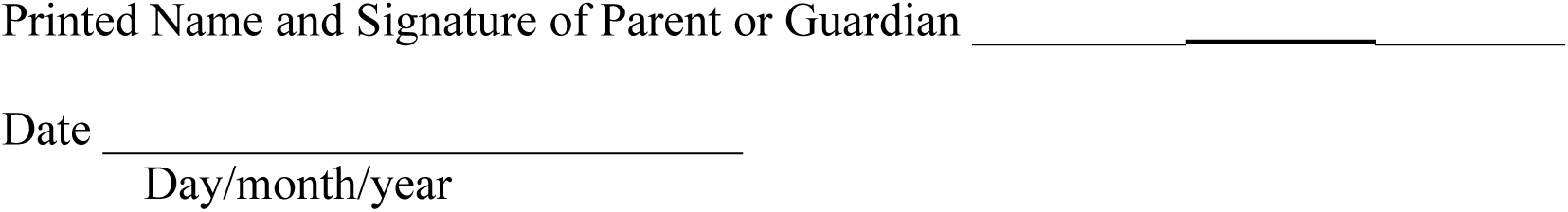

##### If illiterate

I have witnessed the accurate reading of the assent form to the child, and the individual has had the opportunity to ask questions. I confirm that the individual has given consent freely.

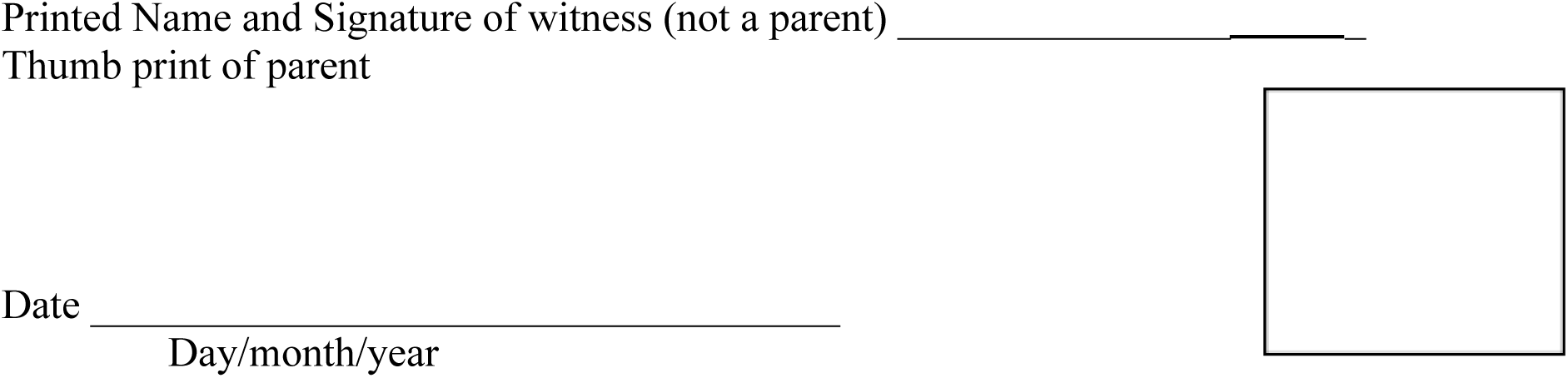

#### Statement by the researcher/person taking consent

I have accurately read or witnessed the accurate reading of the assent form to the parent of the potential participant, and the individual has had the opportunity to ask questions. I confirm that the individual has given assent freely.

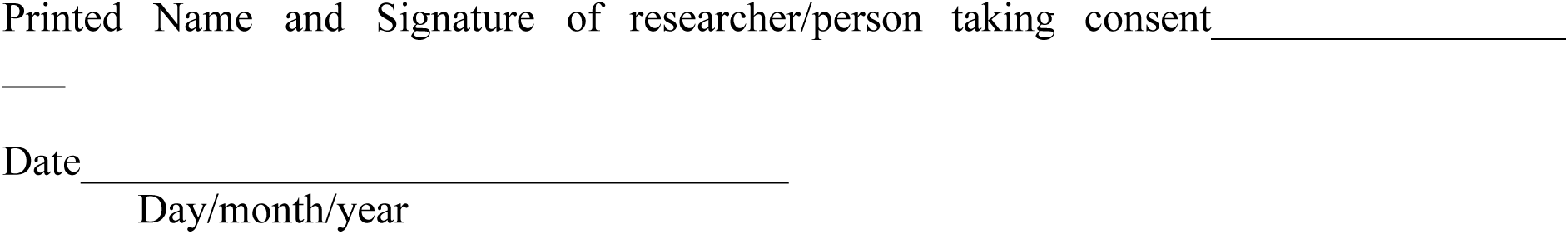

## APPENDIX D GANTT CHART

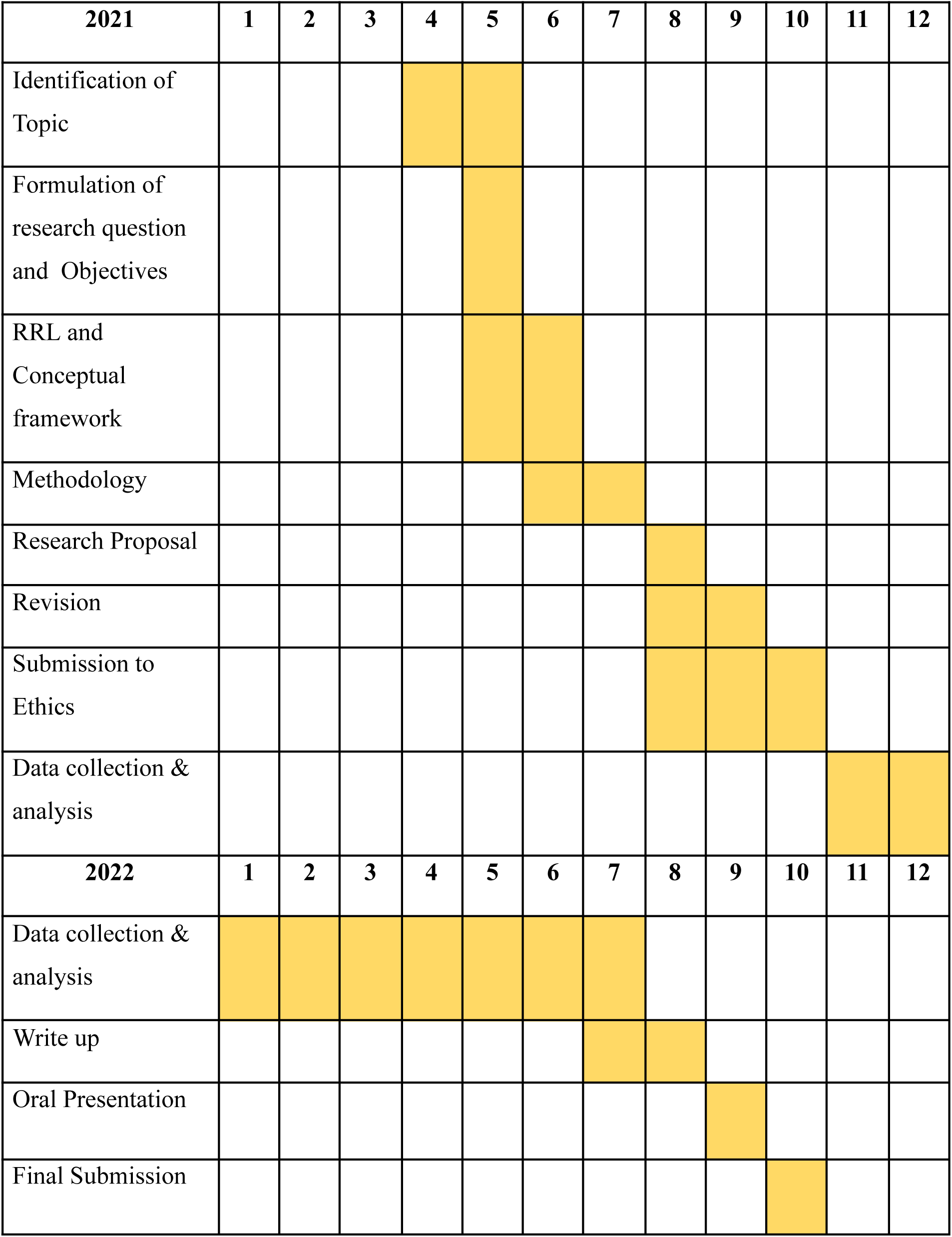

## APPENDIX E BUDGET

| **MATERIALS/ EXPENSES** | **COST** |
| --- | --- |
| Paper | Php 2500.00 |
| Printing | Php 3000.00 |
| Book binding | Php 2000.00 |
| Statistician | Php 7,000.00 |
| **TOTAL** | **Php 14,500** |

## APPENDIX F INSTITUTIONAL ETHICS AND REVIEW BOARD CERTIFICATE OF APPROVAL

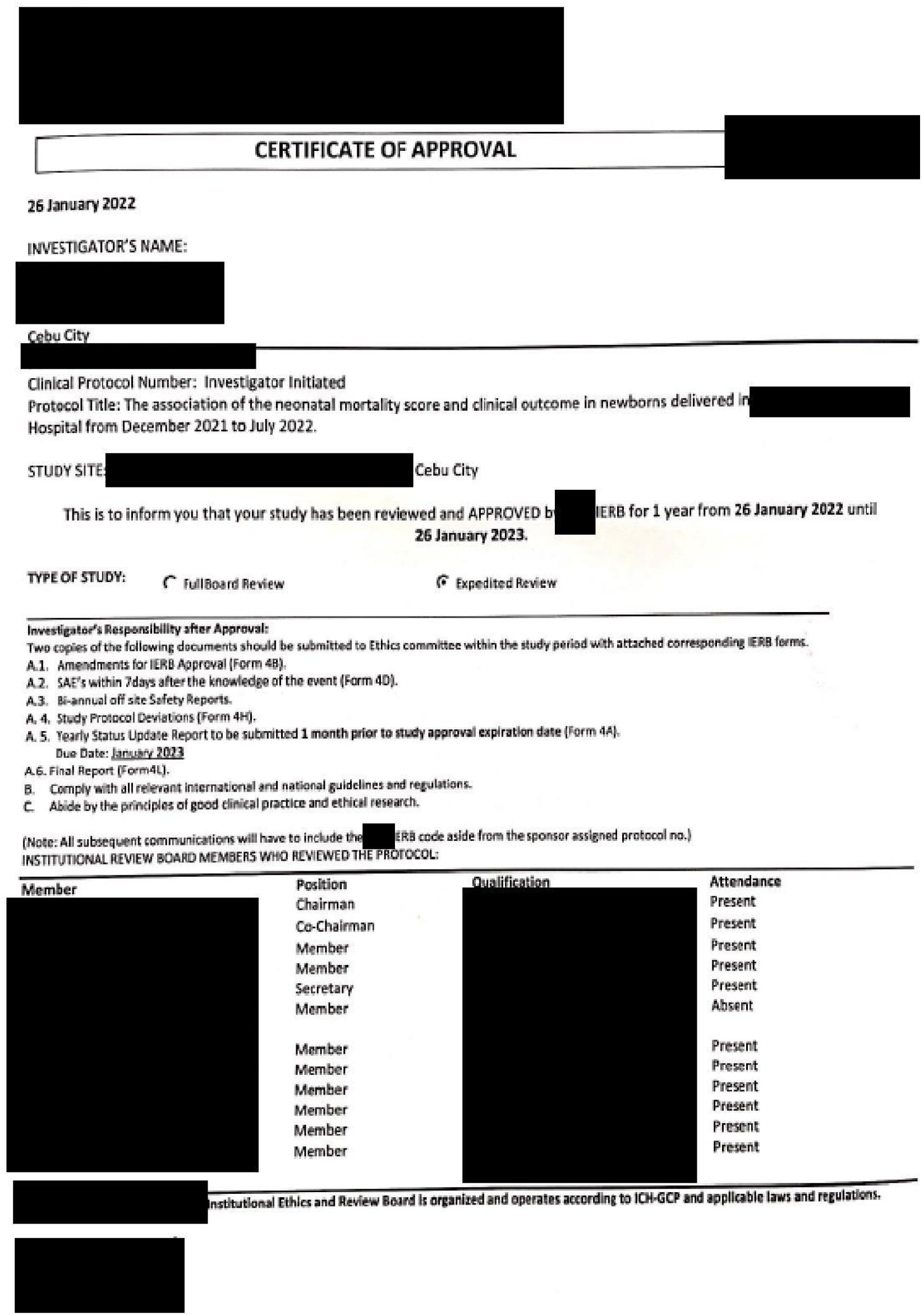

## APPENDIX G PLAGIARISM SCAN REPORT

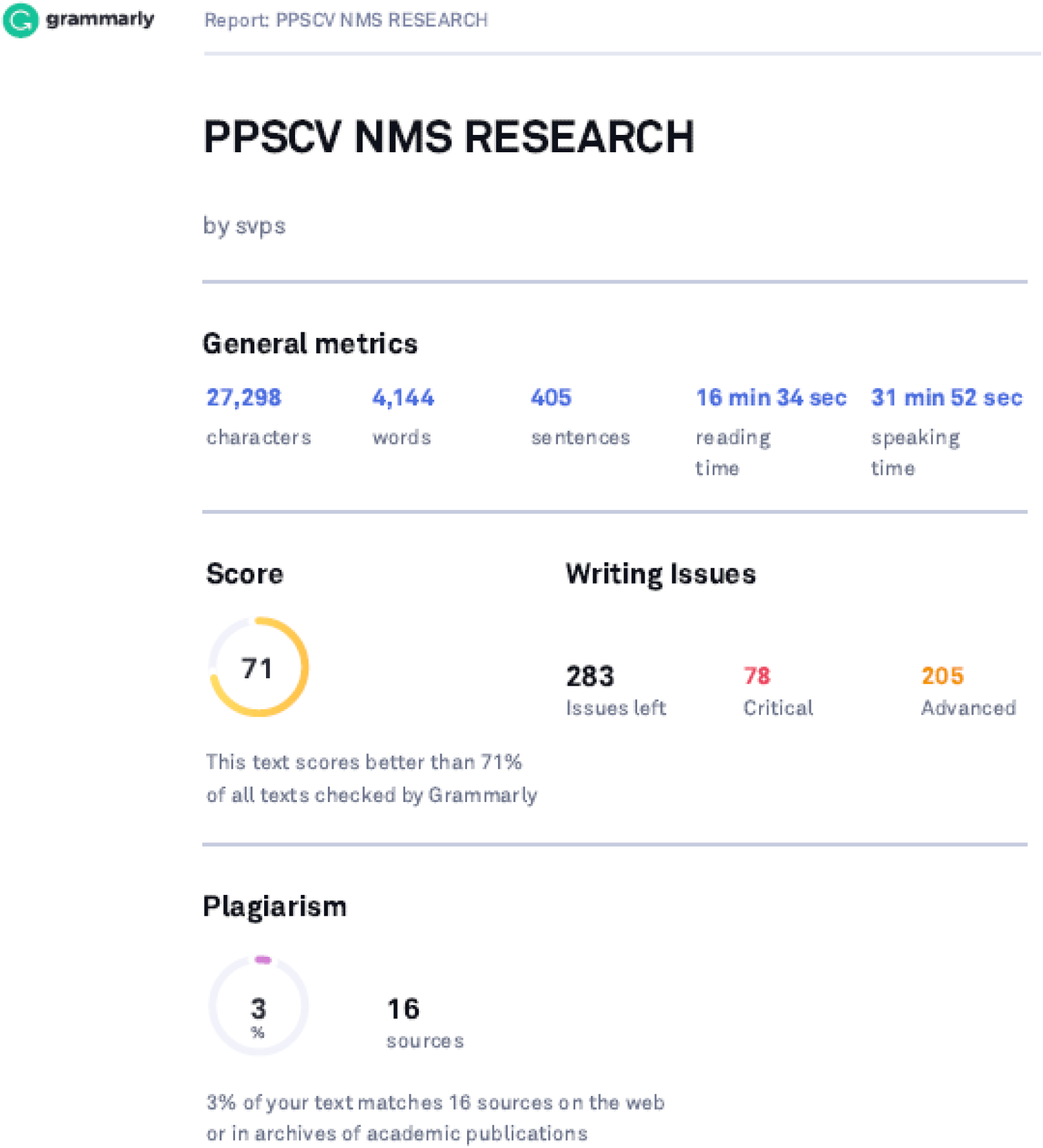

